# EFFECTIVENESS OF A TAILORED SLEEP EDUCATION PROGRAM TO IMPROVE SLEEP AND ITS IMPACT ON ACADEMIC PERFORMANCE IN MULTISPORT YOUTH ATHLETES

**DOI:** 10.1101/2024.10.17.24315671

**Authors:** Ana Merayo, Gil Rodas, Oscar Sans, Alex Iranzo, Lluis Capdevila

**Affiliations:** Medical Department of Futbol Club Barcelona (FIFA Medical Center of Excellence) and Barça Innovation, Barcelona, Spain; Sports Medicine Unit, Clínic Hospital and Sant Joan de Déu Hospital, Barcelona, Spain; Faculty of Medicine and Health Sciences, University of Barcelona, Barcelona, Spain; Sport Research Institute, Universitat Autònoma de Barcelona, Barcelona, Spain; Unitat Trastorns del Son. Servei de Neurologia. Hospital Sant Joan de Deu, Barcelona; Sleep Disorders Center, Neurología Department. Hospital Clinic de Barcelona, IDIBAPS, CIBERNED, Spain; Laboratory of Sport Psychology, Department of Basic Psychology, Universitat Autònoma de Barcelona, Barcelona, Spain; 7

**Author notes:** Corresponding author: Ana Merayo García (AM).

**Keywords:** Academic performance, athletes, education program, mood, sleep, sleep quantity, sleep quality, sports. Sleep

## Abstract

**Objective:** *t*o evaluate the effectiveness of a sleep education program among young athletes in enhancing sleep quality and duration, as well as mood and academic performance.

Design: prospective cohort study.

**Methods:** We included 639 players (11% female; mean age of 13.89±3.8 years) of 5 sports disciplines in a professional club were evaluated before and after a sport season, through 4 specific instruments: 1) sleep diaries to estimate nocturnal sleep duration, 2) the Children’s Sleep Disorder Score Scale (SDSC) to assess sleep quality, 3) the Sleep Vitality Scale (SVS) to examine mood, and 4) school records of academic performance. The sleep education program included staff, family and individual sessions. It focused on the promotion of healthy sleep habits.

**Results:** The 16t-25 years-old (y-o) group exhibited an increase in nocturnal sleep duration (p=0.002), while the 12-15 y-o group showed a decrease (p=0.01). In contrast, the 7-11y-o group exhibited no change. For sleep quality, the 12-15y-o (p < 0.001) and 16-25y-o (p<0.001) groups, while the 7-11y-o group exhibited inferior sleep quality (p 0.001). Regarding mood, the 7-11y-o group showed a significant deterioration (p=0.008), while no changes were observed in the 12-15y-o and 16-25y-o groups. Academic performance exhibited a significant improvement in the 7-11y-o (p=0.001), 12-15y-o (p<0.001), and 16-25y-o (p=0.008) groups.

**Conclusions:** Among athletes aged 12-25y-o, participation in a sleep education program led to improvements in sleep quality and duration, accompanied by enhanced academic performance. However, this intervention did not yield positive effects for athletes between the ages of 7 and 11 years.

## 1. INTRODUCTION

There is mounting evidence that sleep plays pivotal role in optimal athletic performance (1,2). Adequate sleep optimizes physical, cognitive, and emotional well-being (3–4). Studies have demonstrated that partial or total sleep deprivation impairs various aspects of athletic performance, including physical and neurocognitive functions, mood, body composition, and immune function, among others (1,2,5). A growing body of evidence suggests that sleep deprivation may increase the risk of injury in athletes (6, 7). Adequate sleep quantity and quality has also been identified as a recovery strategy due to its physiological and restorative effects (8).

With regard to the physical performance of athletes, there is a growing body of evidence indicating that the sleep deprivation is associated with reduced reaction times in sports such as tennis and basketball (9,10), as well as decreased anaerobic power (10) and endurance (3,5). Furthermore, insufficient sleep has been linked to negative changes in mood and emotional regulation (11). Factors such as stress, frustration, apathy, irritability, and diminished self-confidence can arise due to lack of sleep (12). Sleep deprivation has been associated with impaired learning and executive functions, which are crucial for athletes’ tactical development during training (2,10,13). Other factors, such as demanding training programs, packed schedules, increased travel time, and pre and post-competition anxiety levels, can influence sleep. In the context of sport, it is of paramount importance to maintain an optimal balance between academic pursuits and athletic performance. This is particularly relevant in the case of young athletes, who tend to engage in late-night activities but are required to rise early for academic or training purposes. This can have a detrimental impact on their academic and athletic performance (2).

Additionally, there is evidence suggesting that educational interventions can enhance sleep quality and mood in order to improve academic and sport performance. These interventions aim to mitigate or prevent inadequate sleep hygiene by providing tools to manage habits and psychological variables before, during, and after sleep (14). The interventions focus on reducing hyperactivity, modifying behavior (15) controlling thoughts (16), and establishing protocols to enhance sleep (17). Furthermore, sleep interventions have been demonstrated to be effective in minimizing the negative effects of sleep deprivation and enhancing athletes’ overall well-being (18).

The objective of this study was to assess the effectiveness of a personalized sleep education program throughout a sport season in enhancing sleep quality and quantity, psychological well-being, and academic performance among youth players in elite multisport teams.

## 2. METHODS

### 2.1 Participants

The study included 639 participants (11% were female), with a mean years of 13.89 ± 3.8 (range, 7 to 25) years. All participants were athletes from various sports disciplines affiliated with Futbol Club Barcelona, including soccer, handball, basketball, futsal, and rink hockey. There was a total of 44 teams among the different categories.

The study was conducted during the 2018/19 academic year and sports season. The study was conducted in accordance with the highest ethical standards and data protection policies, overseen by the club’s legal department. All participants were fully informed about the study’s details and provided their informed consent to participate. In cases where athletes were under the age of 18 years, legal guardians also signed the informed consent forms. The study protocol (AS-2019-01) was approved by the Scientific Research Ethics Committee (CEIC) of the International University of Catalonia. The protocol version was 1.0, dated January 21, 2019.

### 2.2 Instruments

#### 2.2.1 Sleep Diary

The quantity of sleep was evaluated by completing a daily sleep diary each morning upon awakening (19). This variable well be referred to as "sleep duration". Participants recorded their estimated sleep onset, instances of nocturnal awakenings (if any), and the time and reason for awakening. The hours of sleep were calculated as the sum from bedtime to wake time, subtracting nighttime awakenings and adding the duration of daytime naps. The data were collected over 5 consecutive nights (Monday to Friday, the measure the typical week of the athletes, excluding days without school exams and 4 weekly training sessions), beginning simultaneously with other questionnaires. To determine whether the sleep duration of athletes is optimal or not, we followed the recommendations provided by The National Sleep Foundation (NSF) (20). Coaches or captains asked participants to regularly fill out the diary. For statistical analysis, sleep hours were converted to a decimal scale.

#### 2.2.2 Sleep Disturbance Scale in Children (SDSC)

This scale was utilized to assess the sleep quality of participants, with a particular focus on sleep behavior and disturbances (21). It encompasses 26 items, grouped into 6 factors, and was completed with the assistance of legal guardians when it was necessary, in accordance with the original study protocol (21).

Participants and guardians responded to statements pertaining to specific sleep-related behaviors using a Likert-5 scale (1 = Never; 5 = Always; total score range: 26-130). A score of 39 points or above indicates the presence of some degree of sleep disturbance. While the SDSC is typically employed in the assessment of individuals under the age of 18, it was utilized in the evaluation of athletes between the ages of 19 and 25, as there are no discernible clinical scales that differentiate between adolescents and young adults.

#### 2.2.3 Subjective Vitality Scale (SVS)

This is a validated tool for assessing dynamic well-being and mood (22). The SVS consisted of 7 items that measured enthusiasm, liveliness, and energy levels. Participants responded on a Likert-7 scale ranging from 1 (completely false) to 7 (completely true). The total score is obtained by adding the item scores, with the highest score beings 49, indicating greater vitality.

#### 2.2.4 Academic performance assessment

The academic grades for the first (December) and last (June) quarters of the academic year were provided by the participants or their families and recorded for analysis. These grades were obtained from the Spanish educational system. The number of academic subjects and the proportion (%) of subjects passed (grades above 5-10) were documented. Participants were notified of their school grades via letters, in accordance with their signed informed consent.

### 2.3 Procedure

The SDSC and SVS scales, along with the sleep diary were administered at the outset of the season (pre-intervention: PRE) and again at the conclusion of the season (post-intervention: POS), following sleep education sessions in which all players had participated. The instruments were initially completed digitally through an online survey platform by the athletes at the start of the season. The SDSC and SVS took approximately 20 to 30 minutes to complete. All participants completed the instruments during the training sessions. To ensure the collection of more precise and truthful information, legal guardians of players under the age of 12 assisted in completing the instruments alongside the participants. A coach and a member of the research team were present during the instruments implementation process to address any queries. All participants completed the tools on the same day, which was a working day of the week, specifically Monday, and they were away from the weekend match. It was ensured that players filled the tools when they did not have any exams at school. Furthermore, participants were required to maintain a sleep diary for the subsequent 4 days, resulting in a total of 5 entries per participant from Monday to Friday. Each player was required to attend a minimum of 4 sports training sessions during the week. In terms of academic performance, we collected data on the players’ grades from the players and their families during the last months of the season. This data provided the percentage of subjects passed by each athlete throughout the academic course.

#### 2.3.1 Sleep education program

The sleep education program was based on a pre-post-intervention design. It was addressed to players and their sports staff, as well as families, as detailed later. Group educational sessions on promoting healthy sleep practices were held for the players’ teams, technical staff, and families. In addition, individual counseling sessions were held with each player. A total of 44 educational group sessions were conducted, 1 for each team, from the category with younger players to the category of the affiliate teams for all sports sections. These sessions, attended by players and technical staff (including coaches, doctors, psychologists, and physiotherapists), emphasized essential aspects of sleep hygiene, focusing on promoting healthy sleep practices and strategies to improve sleep quality in young players. Additionally, group sessions were conducted for the players’ families, with the objective of providing comprehensive knowledge on proper sleep hygiene practices and recommendations. The educational information covered a range of topics, including an overview of sleep and its functions, the physiological changes that occur during sleep, the recommended hours of sleep based on age, and specific recommendations for travel and daily routines.

In addition to the group sessions, 2 sessions were held for each athlete, addressing specific aspects of sleep hygiene relevant to their individual needs. The sessions were tailored to the athletes’ schedules, including school and sports schedules and daily routines. All athletes received 2 comprehensive feedback reports, one at the beginning and the other at the end of the season. These report highlighted the quantity and quality of their sleep based on their responses to the sleep diary and the SDSC. The objective of these reports for the athletes was to awareness of their sleep duration and quality, and to raise awareness of their current sleep hygiene.

#### 2.3.2 Characteristics of the educational sleep program

The intervention was executed meticulously in 3 distinct phases, thereby ensuring a comprehensive approach to the promotion of optimal sleep habits in young athletes.

##### 2.3.2.1 Phase I: Pre-test (The initial 3 months of the season)

In the initial phase of the study, coaches, players, and families were provided with comprehensive information and instructions regarding the program’s nature and its objectives. Participants were informed about the study’s purpose and provided with instructions on how to electronically complete the tools. All participants and their families (in the case of minors) were provided with a detailed report on the results of this initial phase, which included information on their sleep duration and quality, as well as recommendations for improving their current sleep hygiene.

##### 2.3.2.2 Phase II: Intervention Program (4 months)

The objective of this phase was to educate athletes on the principles of optimal sleep hygiene, with the aim of improving their sleep quality and duration. The program comprised 3 distinct types of sessions:

###### 2.3.2.2.1 Sleep education session for athletes and staff

These group sessions were conducted for all athletes of the same sport category together with their technical staff (coach, doctor, psychologist and physiotherapist), with a total of 44 teams in different categories. All participants attended a session led by a board-certified pediatrician sleep specialist. This 25-30-minute educational session was followed by a 10-15-minute questions and answers period. A total duration of 35-45 minutes was proposed to maintain the maximum attention of the players and to allow for extra time to resolve any doubts that the participants may have on the sleep program. The subject was the same for all groups and the main topics covered included understanding the functions of sleep, recommended sleep times according to age, the benefits of good sleep quality, and recommendations to improve sleep quality and establish effective sleep routines. The focus was on maintaining to regular sleep schedules, creating a quiet, dark, and cool sleep environment, avoiding stimulants before bedtime, caring for mattresses and pillows, limiting electronic device use 2 hours before bedtime, avoiding naps exceeding 20 minutes, eating a light dinner 2 hours before bedtime, and cultivating a calm state before going to sleep.

###### 2.3.2.2.2 Sleep educational session for families

A parallel session was conducted exclusively for families, maintaining the same structure of the group session for athletes and staff. The information provided covered technical aspects such as phase delay, feeding, and food metabolism. The session was followed by a 15-minute question-answer segment facilitated by the sleep specialist. The objective was to provide families with information on how to support their athlete’s sleep and nutrition needs. It was recommended that athletes and their families implement the recommendations discussed during these sessions over the subsequent weeks.

###### 2.3.2.2.3 Individual counseling session for athletes

All athletes engaged in 2 individual counseling sessions, 1 for each age group. The focus was to review their daily schedules, teach relaxation and mindfulness techniques, and implement the recommendations elucidated during the educational session. The topics covered in this study included the reduction of electronic devices usage, avoiding dinner 2 hours before bedtime, and maintaining adequate light and temperature in the bedroom. The support materials provided included recommendations, worksheets, sleep and rest guides, and agenda management tools. The sessions were approximately 1 hour in length, and athletes were permitted to consult with their psychologist at any time during the day. These professionals were also present during the educational sessions addressed to family and staff.

##### 2.3.2.3 *Phase III: Post-test* (2 months at the conclusion of the season)

At the conclusion of the study, participants were evaluated using the SDSC and SVS scales, as well as the sleep diary and academic performance evaluations. This phase served as a follow-up to assess the impact of the intervention. All participants and their families (in the case of minors) received a detailed report on the results of this 3ª phase, including a comparative analysis between results from the first and third phases.

### 2.4 Data analysis

The data were presented as means ± standard deviation (SD) for continuous variables, and participant numbers (n) and percentages (%) for categorical variables. Participants were divided into three groups according to their age, namely group 1 comprising athletes between 7 and 11 years, group 2 between 12 and 15, and group 3 between 16 and 25. A mixed Multivariate Analysis of Variance (MANOVA) with a 3*2 design (representing 3 age groups * 2 within-subject PRE-POS situations) was used to explore global differences between pre and post-intervention across all study parameters within each age group. A repeated-measures analysis of variance (ANOVA) was conducted within each age level to assess differences between the PRE and POS intervention periods. Mean differences, along with 95% confidence intervals (CI), and Cohen’s d effect sizes were calculated. All statistical analyses were performed using the SPSS statistical package (IBM, SPSS V25.0, Chicago, IL). Results with p-values <0.05 were considered statistically significant.

## 3. RESULTS

Table 1. Title: General Characteristics of the sample: presents the general characteristics of the sample. It should be noted that only soccer had a contingent of female participants, as detailed in this table.

**Table 1:** General Characteristics of the sample.

| Gender | n | % |
| --- | --- | --- |
| Men | 569 | 89 |
| Women | 70 | 11 |
| Sport | n | % |
| Football (men) | 277 | 43.3 |
| Football (women) | 70 | 11.0 |
| Basketball | 80 | 12.5 |
| Futsal | 65 | 10.2 |
| Hockey | 41 | 6.4 |
| Handball | 106 | 16.6 |
| Age Categories | n | % |
| 7-12 years | 153 | 23.9 |
| 13-15 years | 293 | 45.9 |
| 16-19 years | 193 | 30.2 |
| Academic | n | % |
| Primary School | 159 | 24.8 |
| Secondary School | 286 | 44.7 |
| High School | 112 | 17.5 |
| University | 36 | 5.6 |
| Other | 46 | 7.1 |
| **Overall** | **639** | **100%** |

Data are reported as number (n) and percentage (%) of athletes.

Table 2. Title PRE-POS-intervention differences for the variables of the study by age group: presents the PRE-POS differences for the variables of the study, categorized by the 3 age groups. A mixed MANOVA (3×2) was conducted to examine differences between PRE and POS interventions across age groups. The results demonstrated significant variations in sleep hour evolution between PRE and POS interventions across the 3 age groups (p=0.006).

**Table 2:**
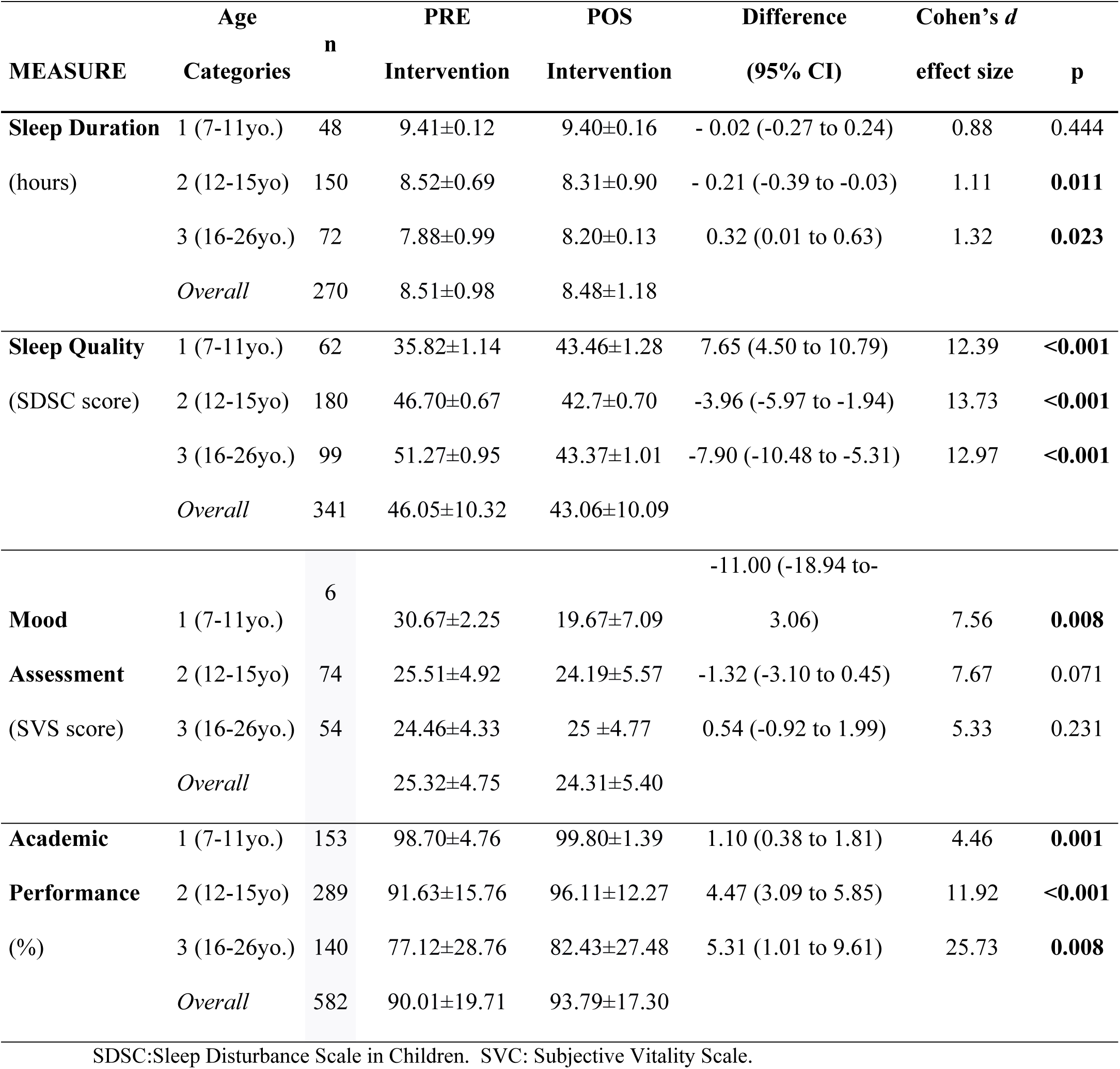
PRE-POS-intervention differences for the variables of the study by age group.

The analysis of sleep duration, as recorded in the sleep diary, revealed distinct patterns among different age groups. Table 2 shows the duration of sleep in a decimal scale. Specifically, the 7-11-year-old group exhibited no changes between PRE and POST intervention periods. In contrast, the group aged 12-15 experienced a significant reduction in sleep hours (12.6 minutes in hexadecimal scale), while the group aged 16-25 showed an increase in sleep duration during the same period (19.2 mines in hexadecimal scale). Fig. 1 illustrates these differences. In particular, the 12-15-year-old group exhibited a significant decrease in sleep hours in comparison to the 7-11-year-old group (p<0.001), which maintained a stable sleep duration. Conversely, the group aged 16-25-year-old group exhibited a significant increase in sleep hours, when compared with the 7-11-year-old group (p<0.001) (see Table 2 and Fig. 1). Fig. 2 presents the percentage of athletes in each group displaying differences in hours slept between POS and PRE-intervention period, as recorded in the Diary. Differences (POS-PRE) are grouped into 15-minute intervals (a negative value indicates that athletes slept less during POS period).

**Figure 1:**
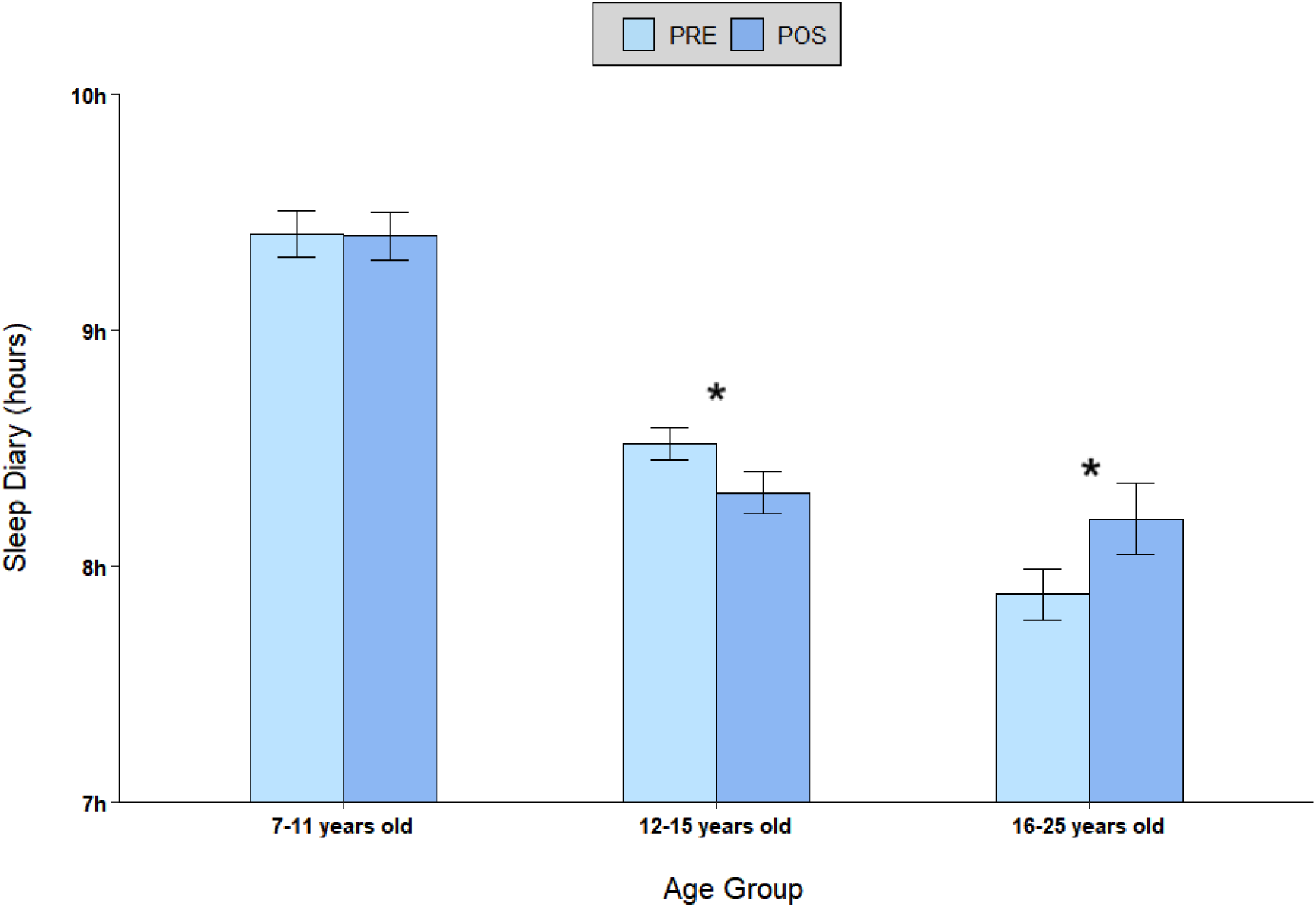
Evolution of sleep hours from the sleep diary between PRE and POS-intervention by age group. * (p<0.001: PRE-POS differences with respect to 7-11 years old group. Bars: Mean and SEM)

**Figure 2:**
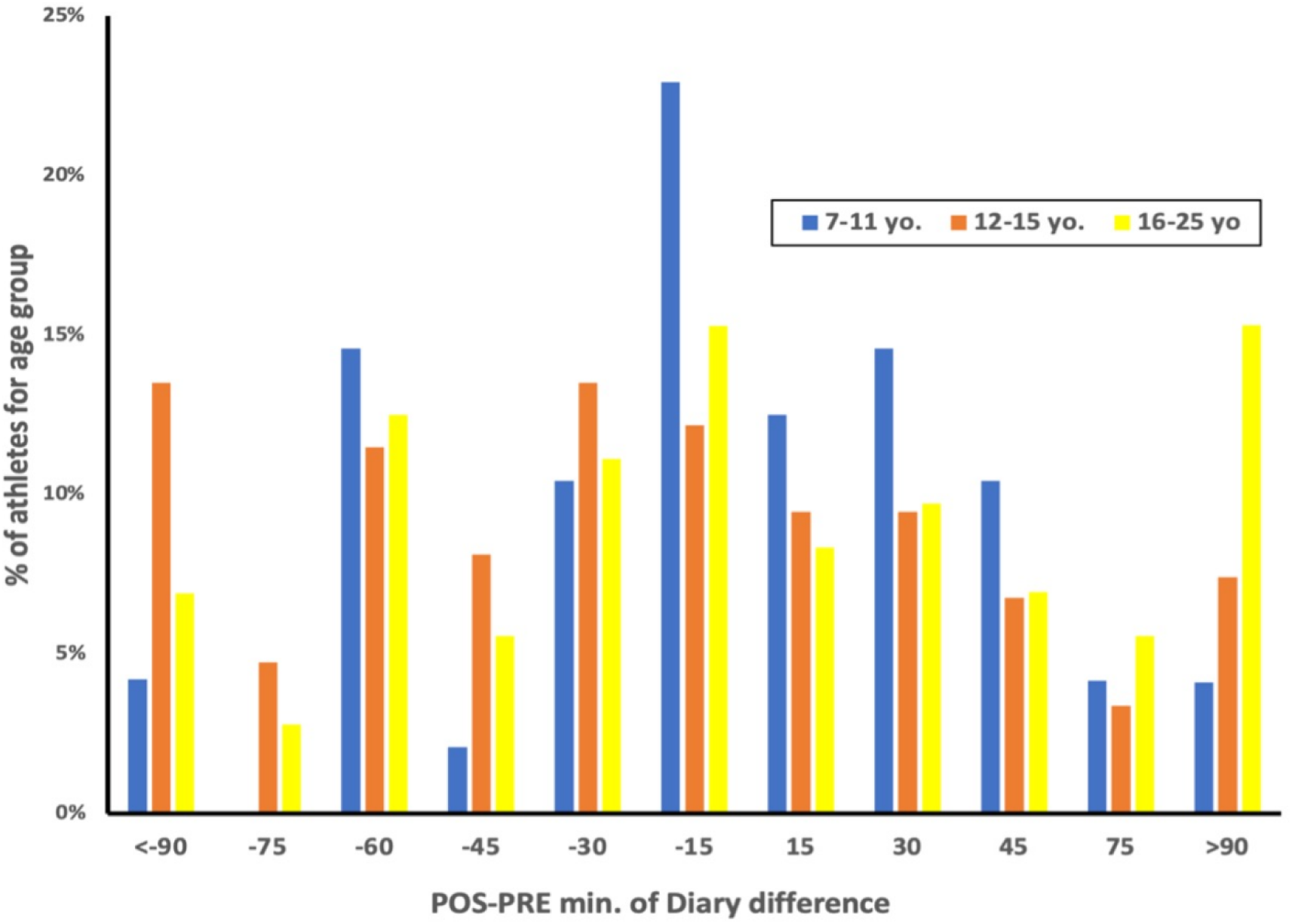
Percentage of athletes by age group according to the variation in minutes slept between POS and PRE-intervention, recorded with the Diary (negative values indicate that athletes have slept less in the POS). 7-11 years old, 12-15 years old and 16-25 years old. Min=Minutes.

When assessing sleep quality was assessed using the SDSC (higher scores indicating poorer sleep quality), the group aged 7-11 displayed a decline in sleep quality. In contrast, both the 12-15 and 16-25-year-old groups exhibited an improvement in sleep quality. Fig. 3 illustrates this distinct behavior of the 7-11-year-old group, which exhibited an increase in the SDSC score (p<.001) compared to the improvement shown by the other 2 age-groups (p<.001). Furthermore, the group aged 16-25 exhibited a more pronounced decline decrease in SDSC score during POS compared to the group aged 12-15 groups (see Table 2 and Fig. 3).

**Figure 3:**
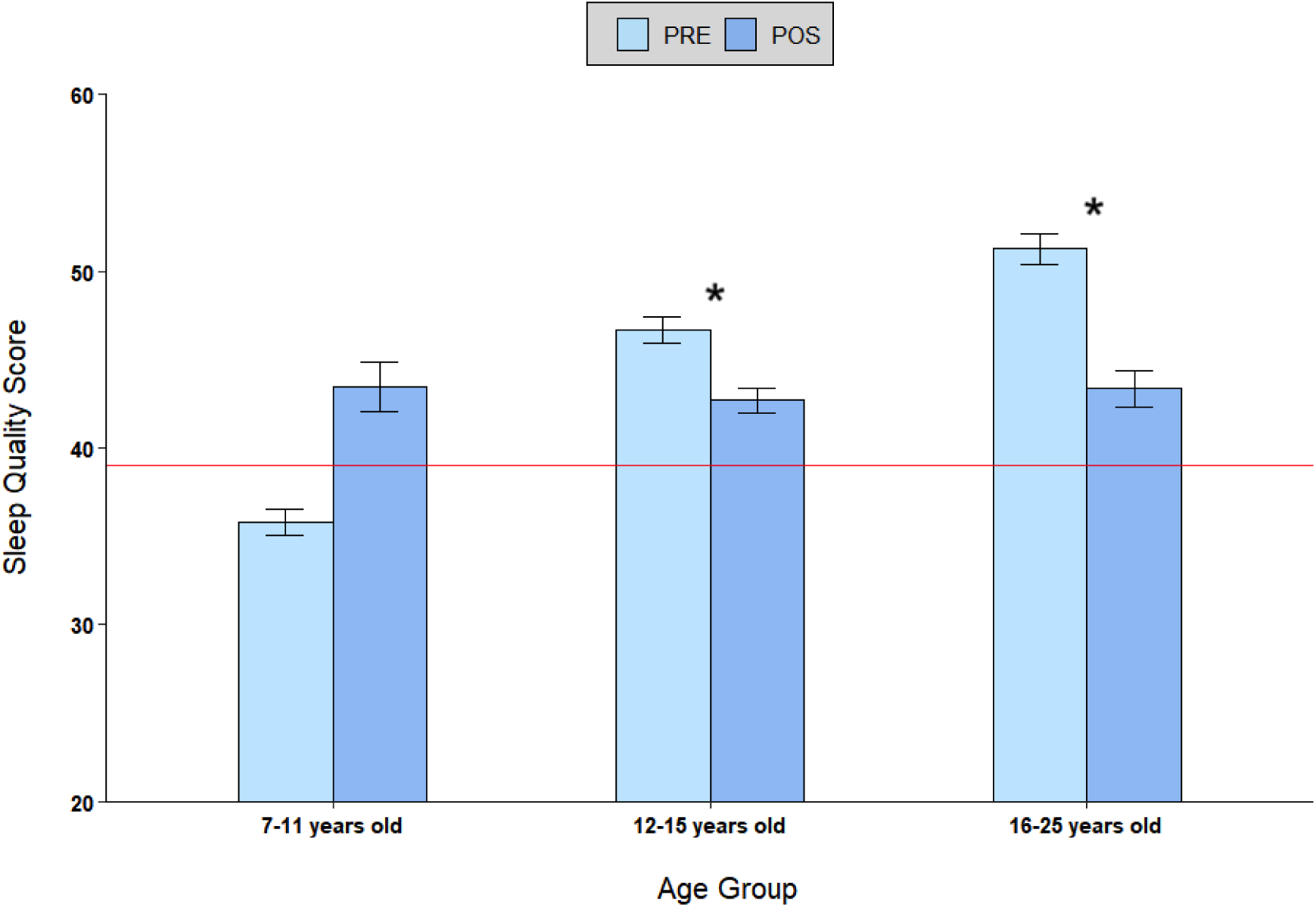
Evolution of SDSC scorings between PRE and POS-intervention by age group. Scores below 39 (red line) indicate higher sleep quality (within the recommended range^38^). * (p<0.001: PRE-POS differences with respect to 7-11 years old group. Bars: Mean and SEM)

Regarding mood, as assessed with the SVS (a higher score indicates a better mood), the 7-11-year-old and the 12–15-year-old groups exhibited a decrease in mood whereas the 16-25-year-old group exhibited no difference. Fig. 4 illustrates that the group aged 7-11 exhibited a lower mood score in POS with respect to the PRE (p=.008), compared to the other two groups (see Table 2 and Fig. 4).

**Figure 4:**
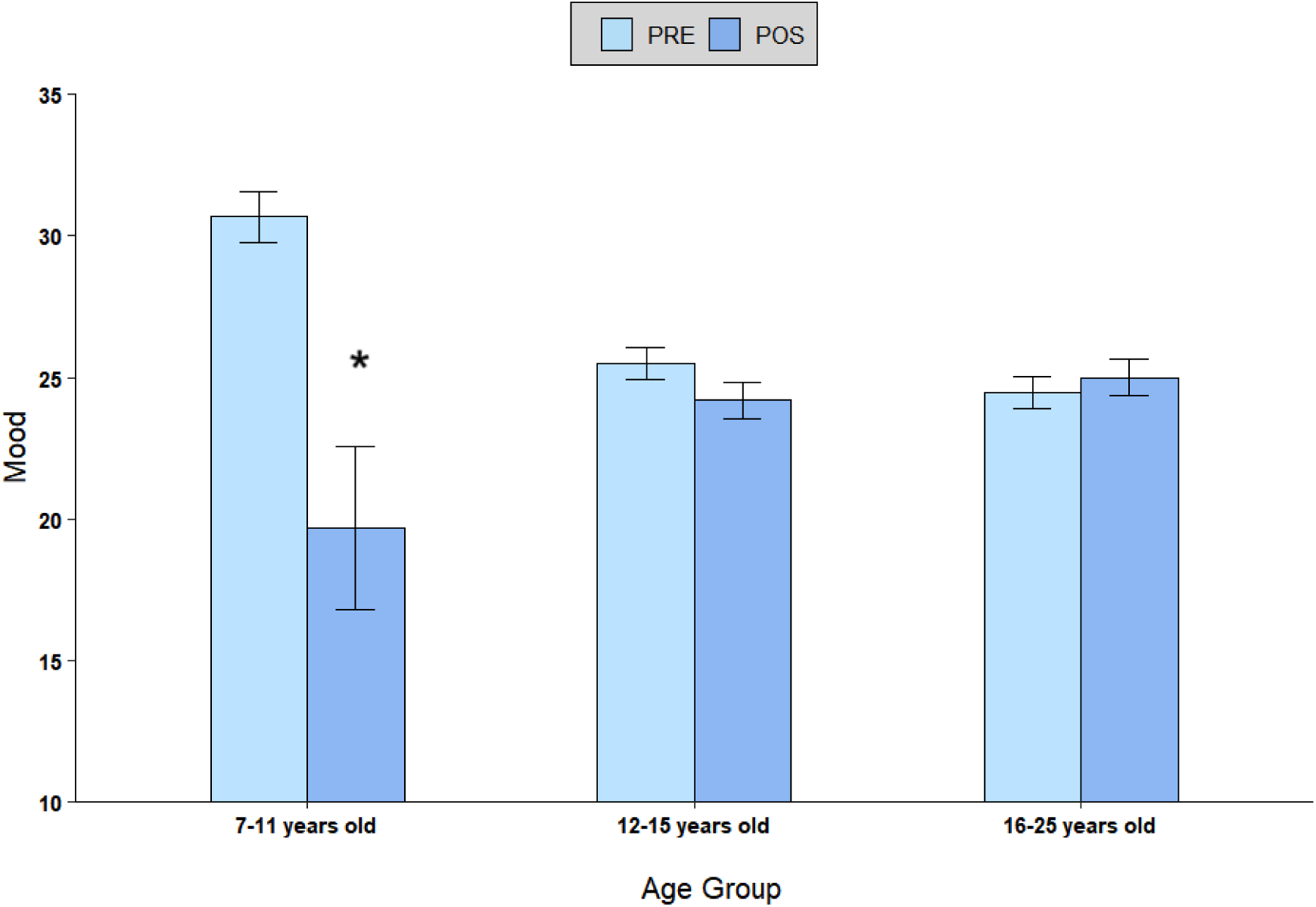
Evolution of mood scorings between PRE and POS-intervention by age group. * (p=0.008: PRE>POS for 7-11 years old group. Bars: Mean and SEM)

For academic performance, all age groups showed a significant increase at the end of the season. Fig. 5 illustrates this change, as measured by the percentage of passed subjects between the PRE and POS periods across different age groups. The entire sample demonstrates an improvement in academic performance during the POS period, increasing from 90% in the PRE period to 93.79% in the POS period. However, the 12-15 (p<.001) and 16-25 (p=.008) age groups exhibited a more pronounced enhancement in academic performance during the POS period in comparison to the 7-11 age group (p=.001) (see Table 2 and Fig. 5). No significant differences were found between genders or among different sports in any of the studied results.

**Figure 5:**
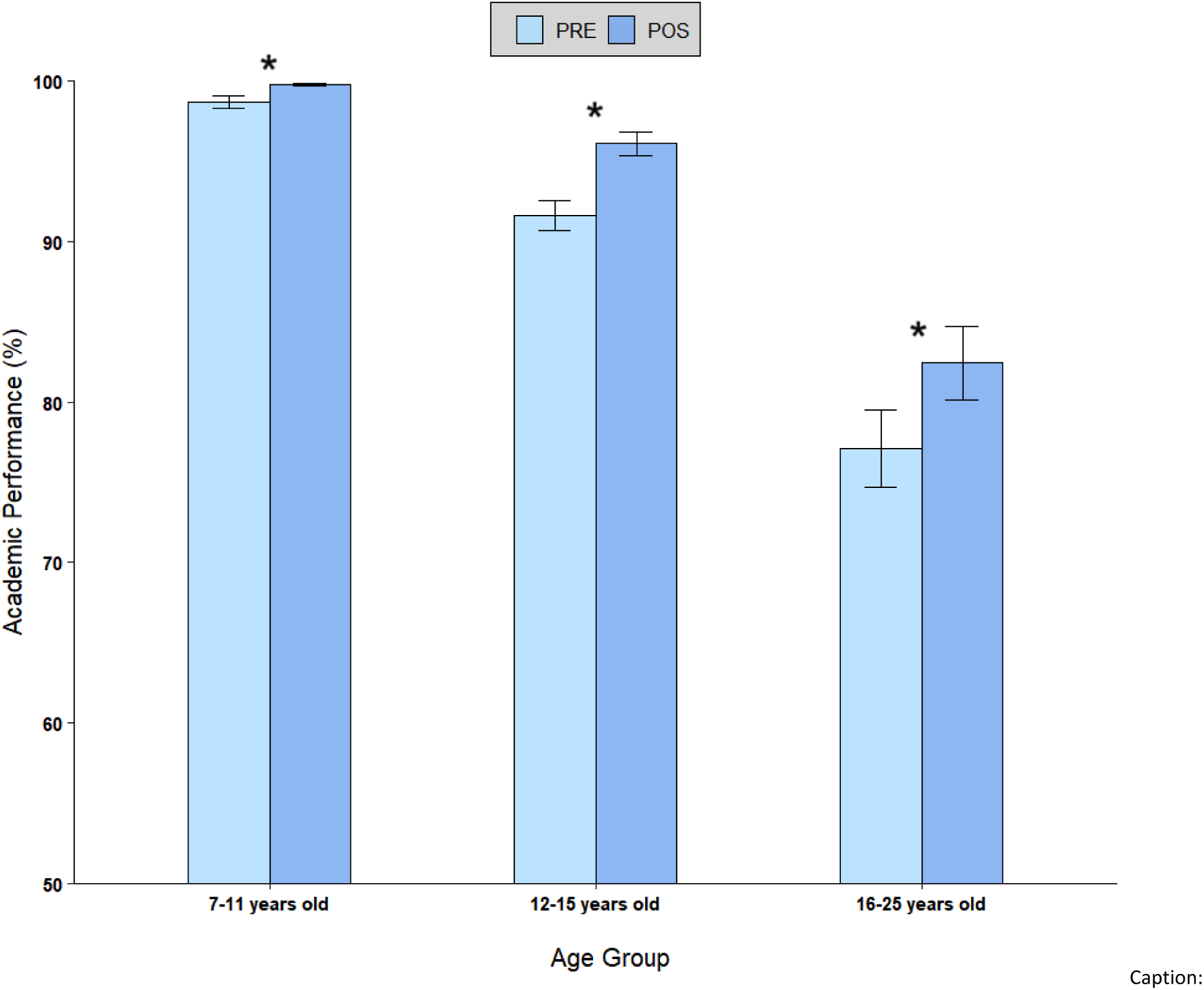
Evolution of percentage of academic passed between PRE and POS-intervention by age group. * (p<0.01: POS>PRE for all age groups. Bars: Mean and SEM)

Table 3 (Change in the scores of the main study variables in the POS compared to the PRE-intervention according to the years age group) presents a summary of the changes in study variables from the PRE to the POS-intervention, with POS serving as a reference point. In summary, for children aged 7 to 11, sleep duration remains consistent in both the PRE and POS-intervention periods. However, in this group there is a decrease in sleep quality and mood scores, and an improvement in academic performance. In the 12-15 age group, there was a decrease in sleep duration and mood during the post-intervention period, but an increase in sleep quality and academic performance. Finally, in the 16-25 age group, there was an increase in sleep duration, sleep quality, and academic performance during the post-intervention period, while mood remained consistent.

**Table 3:** Change in the scores of the main study variables in the POS compared to the PRE-intervention according to the years age group.

| Years Age Group | Quantity Sleep | Quality Sleep | Mood | Academic Results |
| --- | --- | --- | --- | --- |
| 7-11 | = | ↓ | ↓ | ↑ |
| 12-15 | ↓ | ↑ | ↓ | ↑ |
| 16-25 | ↑ | ↑ | = | ↑ |

↑ Improvement; ↓ Getting worse; = Remains the same

## 4. DISCUSSION

The objective of this study was to evaluate the efficacy of a customized sleep education program carried out during a single season in young athletes. The intervention was designed to improve the quality and quantity of sleep, psychological well-being, and academic performance of 639 athletes who participated in an elite multi-sport team. Overall, the intervention was effective across all age groups, with the 16 to 25-year-old group showing the most significant improvement. The intervention resulted in increased sleep duration, improved sleep quality, and enhanced academic performance. Academic performance improved across the entire sample, and sleep quality improved in the intermediate age group of 12 to 15 years. However, the youngest age group (7 to 11 years old) experienced a decline in sleep quality. Mood did not improve in any age group and deteriorated in younger participants.

Sleep duration: The older age group showed the most pronounced increase in sleep duration, affecting approximately 15% of athletes in this group, resulting in an increase of 19.2 minutes in sleep time. This group was the only one averaging less than 8 hours of sleep in the PRE period. In contrast, the intermediate age group experienced a significant decrease in sleep duration, with approximately 13% of athletes in this group sleeping 12.6 minutes less. Most athletes in this group exhibited negative values indicating a reduction in sleep hours during the POS intervention period (see Fig. 2).

This study primarily included soccer players, but the results are applicable to other sports, allowing for generalization. Previous research (23) focused on soccer players and revealed that they generally slept fewer hours than recommended for non-athlete populations of the same age according to the National Sleep Foundation (20). Notably, only the older age group (16-25 years) exhibited an increase in sleep duration post-intervention, with an average of 8.20 hours per night (see Table 2). The average sleep duration was below the recommended levels set forth by the National Sleep Foundation for the 7-11-year and 12-15-year age groups. The latter age group exhibited a reduction in the average duration of sleep, which may be attributed to an increase in mobile phone screen time (24) and late-night training schedules (25)

Sleep quality: The SDSC questionnaire was utilized to assess sleep quality, which demonstrated a significant improvement in the 2 older age groups. This resulted in a reduction in SDSC scores. However, the youngest age group (7 to 11 years old) exhibited a decline in sleep quality post-intervention. Notably, this age group had the highest sleep quality before the intervention, with scores within the recommended range according to the SDSC questionnaire (scores below 39 indicate higher sleep quality (21). The intervention appeared to have equalized sleep quality scores among all groups following the intervention. However, all groups scored above the cutoff point of 39 points for poor sleep quality (see Fig. 3).

In young athletes, the combination of poor sleep quality and insufficient sleep hours indicates a need for specialized attention to improve their sleep-wake habits. Interventions that aim to promote personalized and intensive healthy sleep habits, established at an early age, are crucial for young athletes’ lifestyles (26). The effectiveness of the current intervention may have been insufficient for younger participants, highlighting the need for personalized interventions that are tailored to players and families, considering age, particularly for younger athletes (27). Moreover, older athletes, who benefited more from the intervention, might be more aware of the relationship between recovery behaviors and performance due to their greater mental maturity, underscoring the importance of managing their sleep habits (28)

Academic performance: All athletes, regardless of age group, demonstrated an improvement in their pass percentage following the intervention. The improvement was significantly higher in the 2 older age groups, with an approximate 5% increase in the number of subjects passed during the POS intervention period compared to PRE intervention period (see Fig. 5). The observed increase in the effectiveness of sleep quantity and quality with the intervention could be related to the progressive academic challenges associated with age. Nevertheless, it is crucial to acknowledge that the 7-11-year-old cohort exhibited a remarkably high pass percentage in PRE (97.8%), nearly reaching the maximum pass percentage in the POS intervention period (99.8%) (see Table 2 and Fig. 5). Previous studies have demonstrated that sleep deprivation or poor sleep quality can have a detrimental impact on mental acuity, daily neurocognitive abilities, and academic performance in children and adolescents. Furthermore, it can impair daily functioning and learning abilities (29)

Athletes are able to perform well under pressure, even in inadequate conditions. However, it should be noted that constant sleep deprivation can negatively affect performance and that such circumstances can sometimes result in poor performance. It’s evident that continued sleep deprivation cannot sustain consistently good results (30).

Educational training: our findings underscore the significance of interventions to enhance sleep quantity and quality among young athletes across the lifespan (31). Adolescents in particular should be encouraged to alter their sleep habits through the incorporating of personalized strategies (31). Sleep hygiene is important for promoting overall health and should be accompanied by proper nutrition, hydration, mood optimization, and stress management. These habits are essential not only in sports but also in personal and academic life. It is of the utmost importance to cultivate self-regulating and responsible behaviors in young athletes. Parents and sports staff should be involved in promoting and reinforcing these habits (32).

### 4.1 Strengths and limitacions

The study has several strengths. It includes a large sample of young athletes from Futbol Club Barcelona, a Club with a strong tradition of caring for athletes, promoting invisible training and self-care. Additionally, the study was conducted in a supportive environment. Finally, this study is the first to research and intervene within the same club with such a large number of participants. It provides information on the amount and quality of sleep in a wide and varied sample of athletes, which had not been reported previously.

It should be noted that the study has certain limitations. The study design was based on specific scales and self-report data to collect information about sleep quality and quantity. No objective instruments were used to assess sleep such as the use of mHealth wearable actigraphy and/or polysomnography. The use of actigraphy would have provided data regarding circadian variations between the 3 age groups (33). To control for possible bias, responses from both the group and their parents were included. Another limitation was the duration of the intervention, which was limited to a single season. Extending the intervention period could have revealed additional effects in the study population. Additionally, evaluating sleep data over a limited number of days across 1 season was a limitation. Finally, there is a gender bias as in our study women represented only 11% of the cohort and all women played soccer.

### 4.2 Practical Applications and Future Research

The results of this study could lead to the development of new practical applications and preventative measures not only in the sports population but also in the general population, including early ages and adults. Educational interventions promoting healthy sleep and lifestyle habits could be implemented as a routine among the athletes and their clubs. Dissemination and training on sleep pose a significant challenge. It is important to inform and educate athletes. However, the challenge lies in training the educators and ensuring that families, athletes, and coaches/sports staff receive the same teaching regarding the implementation of adequate sleep habits. The researchers of this study aim to investigate a personalized intervention in sleep hygiene to achieve better results, and explore the relationship between the quantity and quality of sleep, school performance, and mood.

## 5. CONCLUSION

In conclusion, the results of our study indicate that:

- The 2 older age groups, particularly the 16 to 25-year-old group, exhibited significant improvements in both sleep quantity and quality following participation in a sleep educational program.
- The entire cohort demonstrated enhanced academic performance while the mood remained stable across all age groups.
- The intervention improved sleep quality for all groups, with the exception of the youngest group, aged 7 to 11 years old, who experienced a decline.

## Data Availability

The data underlying the results presented in the study are available from Medical center FCB

## Acknowledgments

We extend our gratitude to all FC Barcelona players and staff for their cooperation during the data collection phase. Special thanks to Veronica Tutte and Mario Reyes for their valuable insights and support during the writing of this article. This work was supported from Grants PID2019-107473RB-C21, funded by the Spanish Government (MCIN/AEI/ 10.13039/501100011033), and 2021SGR-00806, funded by the Catalonian Government.

## Author’s contributions

AM conceived the study, programmed the intervention, extracted the data, wrote the materials and worksheets and the final manuscript.

OS conducted the educational program interventions with families and teams. LC conducted the analyses, advised on the analysis, and drafted the manuscript. GR advised on variables All authors have contributed to the revision of the final article, have read and approved the final version of the manuscript and consented to publication, and agree with the order of presentation of the authors.

## Data Availability Statement

The corresponding author will provide the underlying data for this article upon a reasonable request.

## Formatting of funding sources

This research did not receive any specific grant from funding agencies in the public, commercial, or not-for-profit sectors.

